# Medial Arterial Calcification in a 4,500-Year-Old Egyptian King: A Deep-Time View of a Vascular Calcification Continuum

**DOI:** 10.64898/2025.12.09.25341938

**Authors:** Patrick Eppenberger, Katherine D. Van Schaik, Irka Hajdas, Mahmoud Ibrahim, Anna E. Wood, Frank J. Rühli, Moamen Mohamed Othman

## Abstract

**Background:** Arterial calcification is increasingly recognized as a deep-rooted biological process, observed not only in modern patients but also in ancient human remains. While atherosclerosis has been widely studied, medial arterial calcification (MAC), a distinct entity associated with chronic kidney disease (CKD), diabetes, and aging, remains largely unexplored in early populations. This gap limits our understanding of MAC’s historical prevalence and etiologic drivers. During a paleopathological investigation of a mummified forearm attributed to King Unas (Old Kingdom Egypt), we identified arterial calcifications characteristic of MAC, offering a unique opportunity to examine this pathology in a deep-time context.

**Methods:** In situ planar digital radiographs were obtained at the Egyptian Museum in Cairo using a portable system. Arterial calcifications were evaluated according to diagnostic imaging criteria. Osteometric analysis was used to estimate biological sex and stature. AMS radiocarbon dating of associated cranial fragments confirmed Old Kingdom antiquity.

**Results:** Radiographs revealed continuous, tubular, rail-track calcifications along the radial and ulnar arteries, morphologically characteristic of medial arterial calcification (MAC) and distinct from the patchy intimal calcifications of atherosclerosis.

**Conclusions:** This case represents the earliest radiographic evidence of MAC in ancient human remains and demonstrates that medial and intimal calcification predate metabolic disease contexts. In the absence of contemporary risk factors, chronic parasitic infection, particularly schistosomiasis endemic in ancient Egypt, emerges as most plausible contributor to CKD and subsequent MAC. These findings support an evolutionary framework in which MAC and IAC reflect related osteogenic pathways modulated by systemic stressors and vascular bed-specific responses. The case of King Unas thus contributes a rare deep-time data point that broadens the narrative of ancient vascular disease beyond atherosclerosis and underscores the value of integrating paleopathology with modern vascular biology to refine models of arterial calcification.

## Introduction

The progressive calcification of blood vessels is a pathological feature of cardiovascular disease, the leading cause of morbidity and mortality worldwide (1). In contemporary clinical practice, intimal arterial calcification (IAC), primarily associated with atherosclerotic plaque, has long been recognized as a central feature of advanced vascular disease (2). Medial arterial calcification (MAC), characterized by osteogenic transformation and mineral deposition within the tunica media, is increasingly identified as a clinically relevant entity distinct from atherosclerosis (3–6). Traditional risk factors for MAC include diabetes mellitus (3, 5, 7), associated chronic kidney disease (CKD) (3, 8–10), aging (6, 11, 12) and certain hereditary disorders (13, 14). These conditions disrupt phosphate homeostasis, promoting vascular smooth muscle cell (VSMC) transdifferentiation and enhancing vascular mineralization (15–17). The frequent co-existence of IAC- and MAC-related processes in PAD patients suggests an etiopathophysiological continuum influenced by metabolic and mechanical stressors (7, 10, 18–20).

A recent commentary by Thomas and colleagues highlighted the possible evolutionary origins of atherosclerosis, noting that calcified atherosclerotic lesions are prevalent in ancient mummies and may stem from genetic predispositions shaped by evolution (21). They propose that genes conferring early-life advantages (via strong immunity or fertility) persisted despite predisposing to atherosclerosis later in life—an example of antagonistic pleiotropy (21). Indeed, CT analyses of hundreds of mummies across ∼4 millennia have revealed evidence of atherosclerosis in ∼38% of individuals, even though most lived short, pre-modern lives (21, 22). This surprising ubiquity suggests that cardiovascular pathology is deeply ingrained in human biology, transcending modern risk factors. However, previous mummy studies have focused almost exclusively on intimal atherosclerotic calcification. The occurrence of medial arterial calcification (MAC) in antiquity remains largely unexplored (23–26).

To address this gap, we examined the mummified forearm of King Unas (circa 2345 BCE, Fifth Dynasty of Egypt) for signs of arterial calcification. Unas’s remains present a unique opportunity at ∼4,500 years old, they offer one of the oldest examinations for vascular calcification. We sought to determine whether MAC could be identified in this specimen and, if so, how it informs an emerging concept of a vascular calcification continuum from intimal to medial forms.

We situate this case within a paleoepidemiological framework, examining whether ancient exposures such as parasitic infection or nutritional stress could have contributed to arterial calcification independent of lipid-driven atherogenesis. Our findings prompt a re-evaluation of arterial calcification mechanisms, raising the possibility that non-classical stressors in antiquity, particularly chronic infection and associated renal impairment, produced calcific morphologies resembling those seen in modern CKD and systemic disease (27, 28). Infectious disease-related renal dysfunction, nutritional stress, and other systemic factors might have historically converged to produce vascular calcification patterns akin to those seen in present-day high-risk cohorts (18, 19). In this context, we also revisit whether IAC and MAC represent distinct disease processes, or rather reflect positions along a broader, biologically evolved continuum of vascular mineralization (20, 29). By situating a case of MAC in deep antiquity, our study bridges paleopathology and modern cardiovascular biomedicine, providing a long-term perspective on vascular disease. This approach allows us to explore whether arterial calcification phenotypes like MAC are conserved features of human aging and disease, thus expanding the evolutionary narrative recently highlighted in *Circulation* (21).

## Methods

### Specimens and archaeological context

The focal specimen of this study is a mummified left forearm fragment extending to the partially preserved hand, currently housed at the Egyptian Museum in Cairo, Egypt (Inventory N° TR2/12/25/1) (Figure 1). This forearm and two associated cranial fragments form a partial assemblage of human remains attributed to King Unas, the ninth and final ruler of the Fifth Dynasty in Old Kingdom Egypt (∼2375–2345 BCE) (30–32). A thorough historical record traces these remains from their original discovery in 1881 within the burial chamber of the Pyramid of King Unas at Saqqara (33) through multiple relocations, museum exhibits, and brief anthropological examinations (34, 35). Further details on the specimen’s history can be found in supplementary Table S1.

**Figure 1.**
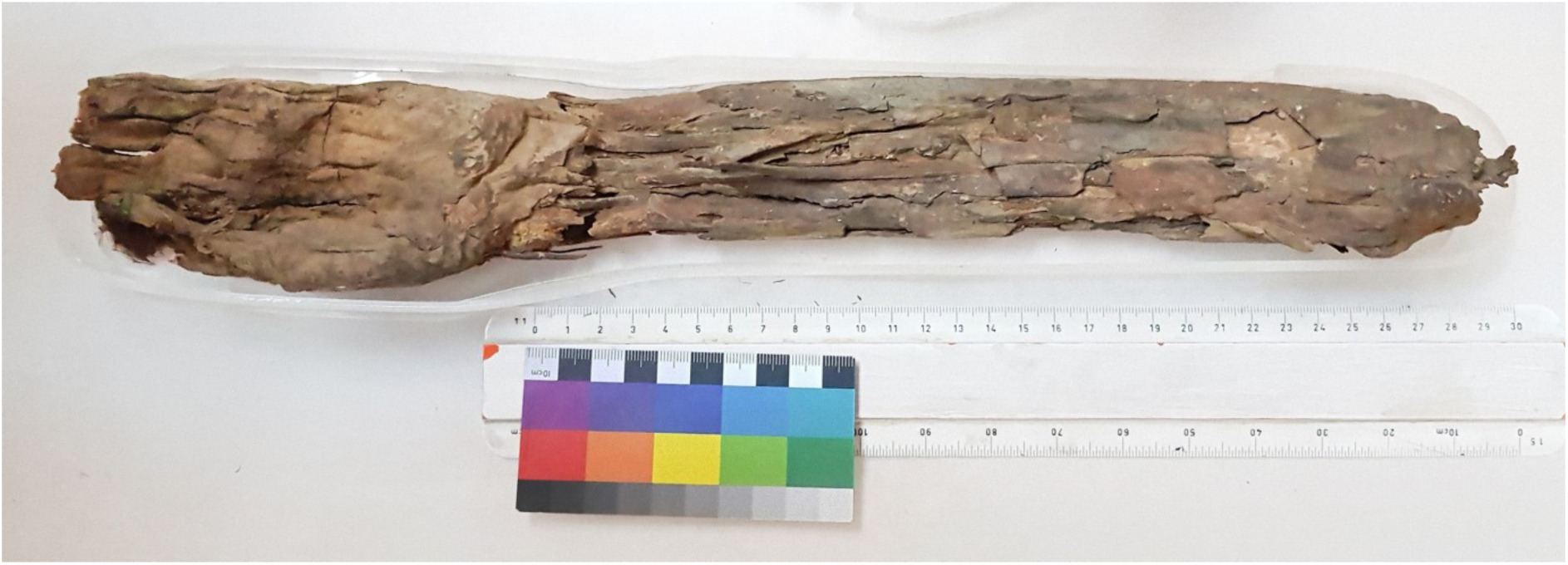
Photograph of the examined mummified left forearm fragment attributed to ancient Egyptian King Unas (Fifth Dynasty, Old Kingdom, ∼2375–2345 BCE).

### Radiocarbon dating

#### Sample collection and preparation

Two small bone samples (1120 and 1290 mg, respectively) were harvested from the cranial fragments in the conservation laboratory of the Egyptian Museum in Cairo (Cairo, Egypt). Contamination was reduced by removing surface encrustations and materials applied during previous museum treatments.

#### Accelerator mass spectrometry (AMS) and calibration

Collagen was pretreated and extracted following established protocols, and radiocarbon measurements were performed via accelerator AMS at the Ion Beam Physics Lab, ETH Zurich (Zurich, Switzerland). Radiocarbon ages were calculated following the methods outlined by Stuiver and Polach (36). The uncalibrated radiocarbon ages for the two samples (ETH-104358 and ETH-104359) were statistically indistinguishable; thus, they were combined to yield a pooled date. The dates were calibrated and combined in OxCal v4.4 (37) using the IntCal20 Northern Hemisphere Calibration Curve (38).

### Imaging

#### Equipment and setup

Due to the fragility of the mummified forearm fragment, planar digital radiographs were obtained on-site at the Egyptian Museum in Cairo (Cairo, Egypt) using a portable digital X-ray system (EXAMION PX 60 HF, Fellbach, Germany). The system consisted of an X-ray generator (maximum output of 3.2 kW, voltage range 40–100 kVp, exposure range 0.4–100 mAs) and a flat panel detector (DR 1417-600 WL, gadolinium oxysulfide scintillator, with a 3072 × 2560 matrix, 140 μm pixel pitch, and 14-bit grayscale). The specimen remained within its plexiglass exhibition mount to prevent handling damage throughout imaging. Radiographs were acquired in two orthogonal projections, dorso-palmar (DP) and lateral (LAT), with the forearm rotated in approximately 90° pronation relative to the elbow due to its mummified state. The source-to-detector distance was set at 150 cm for both projections. The specimen-to-detector distance was kept less than 1 cm, including the plexiglass thickness and a minimal gap between the inner surface of the holder and the specimen.

#### Image processing and analysis

All digital radiographs were stored in Digital Imaging and Communications in Medicine (DICOM) format. Subsequent viewing and preliminary assessments were performed using the open-source Horos (version 3.2.1) software on a 27-inch iMac (AMD Radeon Pro 5-Series GPU) at the Institute of Evolutionary Medicine (University of Zurich, Zurich, Switzerland). Two investigators (PE and KDV) jointly evaluated the images in consensus, examining bone morphology, soft tissue preservation, and radiopaque structures suggestive of arterial calcification. Special attention was given to distinguishing medial versus intimal arterial calcifications based on established radiographic criteria, such as tubular versus patchy calcific densities (39, 40). Additionally, joint surfaces, including the radiocarpal, metacarpophalangeal, and proximal interphalangeal articulations, were assessed for degenerative changes, osteophytes, or morphological alterations indicative of chronic stress or advanced age. Potential taphonomic and embalming-related modifications, defects, or linear fractures were also carefully examined to differentiate postmortem alterations from in vivo pathology (41, 42).

### Anthropological assessment

#### Osteometric measurements

To estimate stature, skeletal robustness, and biological sex, osteometric measurements of the forearm bones were performed directly on the digital radiographs using the internal calibration reference from the X-ray system. Measured parameters included maximum radial and ulnar length and humeral epicondylar width. These measurements were compared to standardized forensic and bioarchaeological reference data (43, 44) to estimate biological sex and living stature.

### Ethical and permitting considerations

All investigative procedures, including radiographic imaging and sampling for radiocarbon dating, were conducted in close collaboration with, or directly by, the Egyptian Museum’s Conservation Department. Permits and approvals complied with Egyptian Antiquities regulations and international ethical guidelines for research involving human remains (ICOM). Due to strict ethical, cultural, and conservation constraints, especially regarding the integrity of the unique forearm fragment, invasive procedures such as histological sampling or molecular analyses were not permissible. Therefore, diagnostic confirmation was necessarily limited to non-invasive radiographic assessments.

## Results

### Radiographic findings

No prior radiographic studies have been reported for this forearm specimen or the associated cranial fragments. Planar digital X-ray images of the mummified left forearm (including the hand and distal humerus fragment) were obtained in two projections, dorso-palmar and lateral. In contrast to standard clinical radiographic projections of the forearm, the limb is seen in approximately 90° pronation due to its mummified state (Figure 2). The radius, ulna, carpals, metacarpals, proximal phalanges, and a portion of the distal humerus are visible. Mummified soft tissues surrounding these bones are also discernible in both projections.

#### Bone and soft tissue observations

A substantial, sharp-edged defect is present near the distal end of the radius and ulna, resulting in an approximately 15 mm gap between the epiphyseal portions (i.e., radial distal end and ulnar styloid) and the diaphyseal shafts. This discontinuity extends partly into the adjacent soft tissues, which remain continuous and well-preserved. Linear, transversely oriented lucencies are visible, crossing the cortices of the diaphysis of the third metacarpal and the metadiaphyses of the fourth and fifth metacarpals. Similar sharp-edged discontinuities appear in the middle phalanges; no distal phalanges are preserved. The trabecular bone structure is demineralized, especially at the metacarpophalangeal joints. Mild osteophytosis of the radial head and subtle joint-space widening of the scapholunate interval suggest some degenerative change but are not pronounced. There is subchondral sclerosis about the triscaphe joint, compatible with degenerative changes. No overt evidence of traumatic lesions or postmortem damage (aside from the noted sharp-edged defects) was detected. The distribution of these defects may reflect taphonomic processes or partial anthropogenic interventions (e.g., embalming, museum preparation).

#### Evidence of arterial calcinosis

In the dorso-palmar view, an elongated, heterogeneously radiodense structure 3-4 mm in width is present between the ulna and radius, following the course where one would anticipate the ulnar artery. A similar elongated structure, also approximately 3-4 mm in width and of comparably increased radiodensity, is projected over the radius, corresponding to the likely course of the radial artery. These two parallel tubular calcifications are likewise identifiable on the lateral projection, partially superimposed upon the radius and ulna. The continuous, rail-tracking morphology of these calcifications, extending throughout the expected arterial trajectory, differs markedly from the patchy, nodular lesions typically associated with IAC at high-flow vascular bifurcations [25,50]. Instead, their elongated, sheet-like pattern aligns with MAC [14,20,28]. While definitive histological confirmation is lacking, these imaging features strongly suggest MAC affecting the forearm arteries.

**Figure 2.**
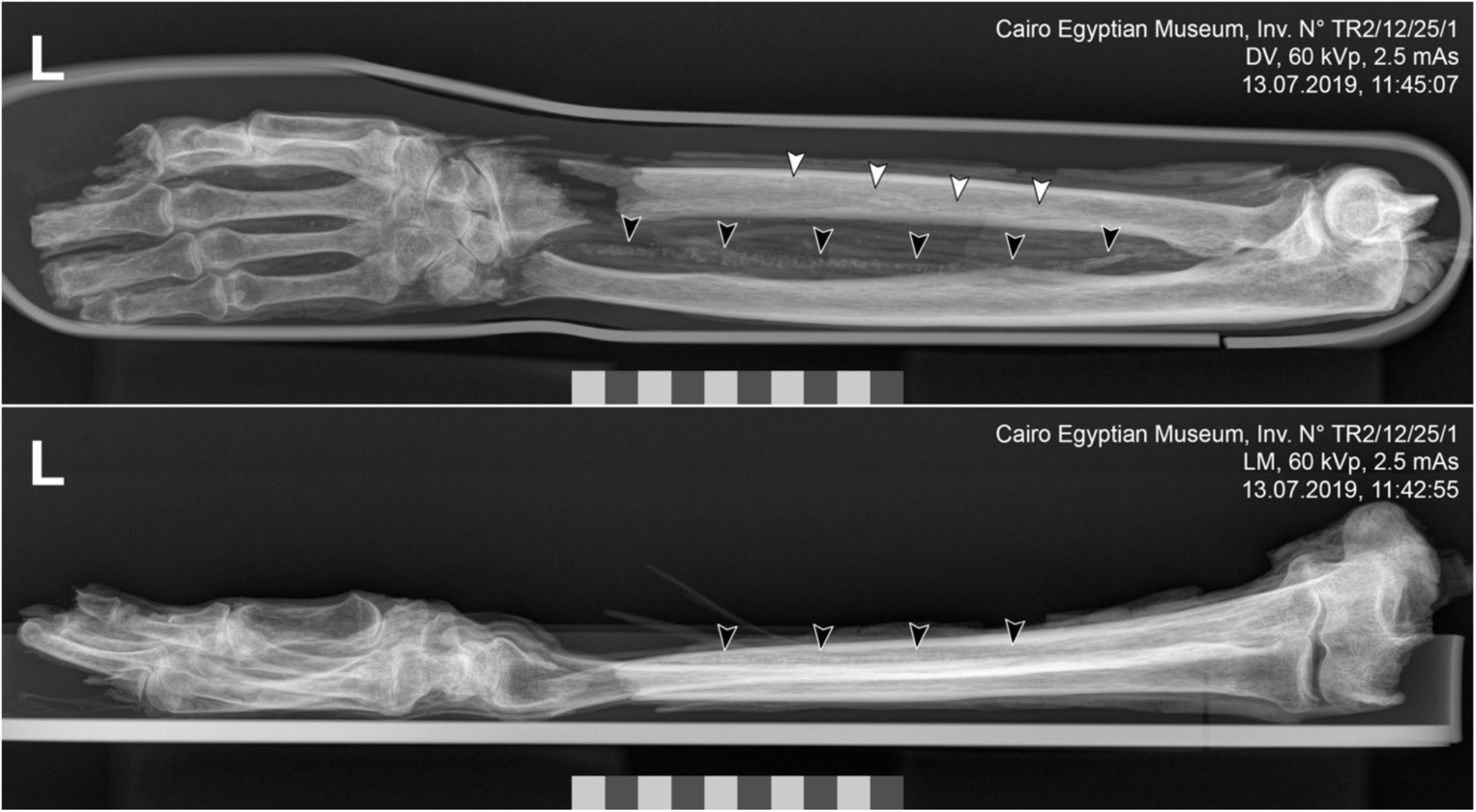
Dorsal-volar (top) and lateral (bottom) radiographs show the radius, ulna, metacarpals, carpals, proximal phalanges, and an attached distal humerus fragment. In the dorsal-volar image, between the ulna and the radius, an elongated structure of heterogeneously increased X-ray density measuring 3-4 mm in width projects over the expected course of the vascular bed of the ulnar artery (black arrowheads). This structure is also identifiable on the lateral image (black arrowheads). Another similar pattern is seen projected onto the radius along the approximate course of the radial artery (white arrowheads).

### Basic anthropological assessment

Osteometric measurements performed on the digital radiographs using the internal calibration reference from the X-ray system included a maximum radial length of 245 mm (including the distal gap), maximum ulnar length of 263 mm (measured to the distal gap margin), and humeral epicondylar width of 60 mm. The humeral epicondylar width is consistent with a male individual based on established forensic standards derived from modern European samples (43). Using ancient Egyptian stature estimation models (44), a radial length of 245 mm equates to an approximate height of 165.6 cm for a male. The cubital, carpal, metacarpal, and phalangeal joints show little degenerative change, suggesting that this individual was not of advanced age at death and/or undertook relatively little physical work with the upper extremities. The demineralization and rarefication of trabecular structure observed in the entire hand suggest osteopenia, which may be related to age, disuse, malnutrition, metabolic disease, or a combination of these factors.

### Radiocarbon dating

No prior radiocarbon dating has been reported for this forearm specimen or the associated cranial fragments. In this study, two samples (ETH-104358 and ETH-104359) taken from the cranial fragments were subjected to radiocarbon (^14^C) dating. Each sample produced nearly concordant uncalibrated radiocarbon ages of 3992 ± 21 BP (ETH-104358) and 3994 ± 21 BP (ETH-104359). A combined calibration placed these remains between 2571-2467 calBCE (95.4% confidence) (Figure 3). This range is older than the historically accepted reign of King Unas (mid-24th century BCE).

**Figure 3.**
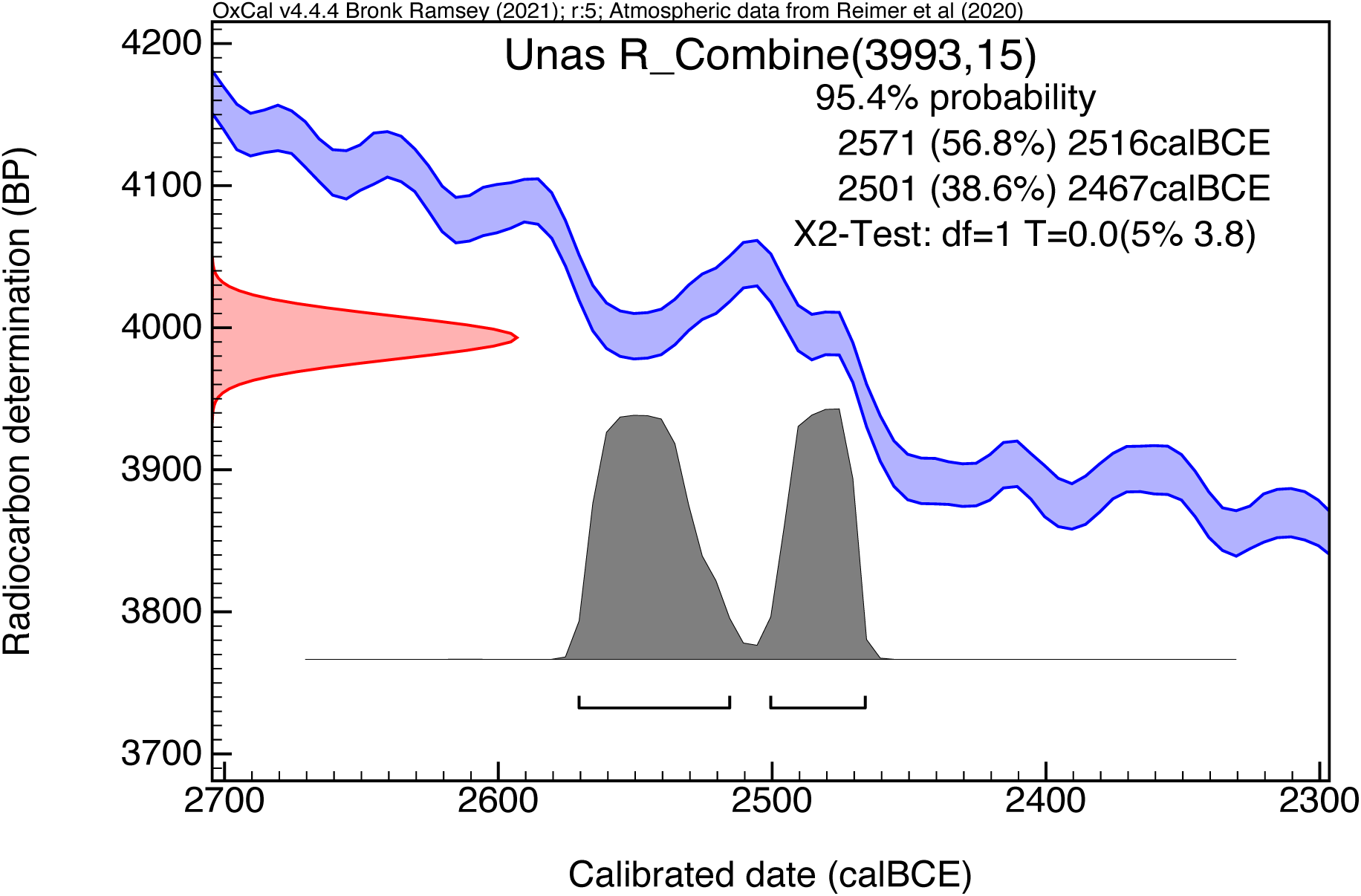
Radiocarbon dating of cranial fragments. Combined radiocarbon date for samples ETH-104358 and ETH-104359. Calibration and combination were completed using the IntCal20 Northern Hemisphere Calibration Curve (Reimer et al., 2020) in OxCal v4.4 (Bronk Ramsey, 2009).

## Discussion

Our findings reinforce the emerging view that cardiovascular disease processes have deep evolutionary roots, as recently highlighted by Thomas et al. (21). The discovery of MAC in a 4,500-year-old mummy demonstrates that the propensity for arterial calcification – beyond just atherosclerotic plaques – has long been part of the human condition. This aligns with the idea that genetic and biological factors predisposing to vascular calcification have been conserved over millennia.

This study presents radiographic evidence suggesting elongated peripheral arterial calcinosis in an ancient mummified left forearm (∼4400 years old), potentially belonging to King Unas of the Fifth Dynasty in Old Kingdom Egypt. The observed arterial calcifications project over the course of the radial and ulnar arteries in dorso-palmar and lateral radiographic views. Their continuous, tubular, rail-track-like morphology is consistent with MAC, as opposed to the irregular patterns typical of IAC reported in previous mummy studies (23, 24, 26). This places the forearm specimen among the earliest known examples of probable MAC in human remains, extending evidence that complex forms of arterial calcification were not limited to modern or post-industrial societies (23–26). Such findings reinforce that vascular calcification may arise under various metabolic or infectious stressors, even in historical contexts lacking many of today’s recognized cardiovascular risk factors.

Importantly, our study complements earlier observations of atherosclerosis in mummies (21, 23–26). While prior research documented intimal calcifications (consistent with atherosclerotic plaque) in ancient populations, we report a case of medial calcification from the same era. The coexistence of these two forms of vascular calcification in antiquity supports the concept of a vascular calcification continuum (20, 23). Rather than viewing MAC and intimal arterial calcification (IAC) as entirely separate entities, our findings align with the perspective of context-dependent manifestations of a shared pathophysiological spectrum.

The osteogenic mechanisms driving vascular calcification (such as bone-like matrix deposition by vascular cells) are known to operate in the medial and intimal layers. Our finding of MAC in a pre-modern context suggests that these fundamental mechanisms were active long before industrial risk factors emerged. In evolutionary terms, this persistence implies a conserved biological response: human arteries have long been capable of mineralization in response to chronic stressors. This idea dovetails with the antagonistic pleiotropy hypothesis raised by Thomas et al., wherein traits beneficial for early survival (e.g., robust inflammatory responses) may have the late-life downside of promoting calcific disease (21). The presence of MAC in an ancient individual supports the notion that the liability of vascular calcification is ancient, potentially tolerated in our species due to earlier-life benefits or neutral survival impact.

Our case also highlights environmental factors in antiquity that could drive vascular calcification. Notably, King Unas lived in a time and region where chronic infections such as schistosomiasis were endemic. We hypothesize that infection-induced chronic kidney disease (CKD) and resultant phosphate imbalance could have promoted MAC in this individual. This scenario suggests that, beyond genetic predisposition, long-standing environmental pressures (e.g., infectious disease burden) contributed to arterial calcification in historical populations. Such factors are less emphasized in modern contexts but were likely significant in the past, indicating that the evolutionary story of vascular disease is multifaceted – involving inherited traits and historical exposures.

### Parallels with Modern MAC and Potential Etiologies

Medial arterial calcification (MAC) is associated with increased arterial stiffness, elevated pulse pressure, and left ventricular afterload. In modern contexts, it is primarily linked to CKD (8), where disruptions in phosphate metabolism, such as hyperphosphatemia and impaired excretion, drive vascular smooth muscle cell (VSMC) osteogenic differentiation, triggering an adaptive but pathological response (12, 16). These pathways, well-characterized in contemporary vascular calcification studies, indicate that MAC is an active, cell-mediated process rather than a passive mineral deposition (15, 45), and despite a prolonged and clinically silent course, ultimately leads to compromised hemodynamics associated with chronic limb-threatening ischemia (46). Radiologically advanced CKD-associated MAC frequently presents with tubular or sheet-like calcifications within the vessel wall, resembling the elongated morphology observed in our forearm specimen (45). Recent studies demonstrate that CKD-related MAC involves an imbalance between pro-calcific (e.g., BMP-2, RUNX2) and anti-calcific (e.g., matrix Gla protein) regulatory factors. This mechanism may have also been triggered by chronic systemic stressors in antiquity (12, 14, 16, 19). While epidemiologic data for CKD in ancient Egypt are lacking, renal pathologies are well documented in Egyptian mummies: abscesses caused by Gram-negative cocci (47, 48), renal calculi (49), nephrosclerosis in a mummy with atherosclerosis (50), and congenital renal atrophy (49). While direct evidence for CKD in ancient populations is scarce, several conditions known to contribute to CKD today, such as chronic infections, metabolic disorders, and dietary factors, warrant consideration in this case.

#### Infectious CKD

Chronic infections remain a significant cause of CKD-related mineral dysregulation. Several pathogens, including tuberculosis, hepatitis C, and schistosomiasis, are implicated in CKD development (51–54). Among these, *Schistosoma haematobium*, endemic to the Nile region for millennia, is particularly relevant. This parasite is a known cause of chronic urinary tract inflammation, renal fibrosis, and subsequent phosphate imbalances, potentially leading to MAC. Schistosomiasis was, until recently, a prevalent cause of chronic renal and bladder pathology in individuals living along the Nile River delta, with a prevalence of up to 30% identified in some villages as recently as 1996; prior rates were as high as 60% in districts of perennial irrigation (53, 55). The ova of these parasites have been identified in the renal tissues of mummies (48, 56), and hematuria, a sequela of schistosomiasis, is reported in the Ebers Papyrus. Given the high prevalence of schistosomiasis in antiquity, infection-driven CKD emerges as a plausible contributor to vascular calcification in this individual.

#### Diabetes Mellitus (DM1/DM2)

While diabetes is a major risk factor for MAC today, its role in this case is less certain. Type 1 diabetes (DM1) existed in antiquity and can accelerate vascular calcification, but survival into adulthood without insulin therapy would have been highly improbable. Though documented symptomatically in the Ebers Papyrus, untreated DM1 would likely have led to fatal metabolic complications before significant vascular calcification could develop. Type 2 diabetes (DM2) remains a theoretical possibility, but its prevalence in ancient populations is unknown, making it a less likely explanation for MAC in this case.

#### Age and Diet

Anthropological assessment suggests the individual was not of advanced age, yet a middle-adult death (potentially after a 15- to 30-year reign) could have allowed sufficient time for degenerative vascular changes. Dietary factors may have also influenced vascular health. Though not fully reconstructed, Old Kingdom diets were rich in cereals, fish, and salt-preserved foods, potentially affecting sodium balance and phosphate metabolism (57). Furthermore, social stratification may have influenced dietary access, with elites consuming more meat, alcohol, or sugar-rich fruits, all of which could modify cardiovascular risk profiles.

#### Hereditary Factors and Endocrine Disorders

Rare genetic conditions, such as *ENPP1* and *NT5E* mutations, can lead to early-onset arterial calcifications. However, their prevalence in historical populations is unknown, and without molecular genetic data, this remains speculative. Similarly, endocrine disorders such as primary hyperparathyroidism, characterized by chronic hypercalcemia and phosphate dysregulation, could contribute to MAC, but there is insufficient evidence to confirm this in our case.

#### Summary of Likely Etiologies

Among the potential contributors, infection-driven CKD (particularly from schistosomiasis) and age-related degenerative changes appear to be the most plausible explanations for MAC in this individual. Diabetes and hereditary disorders are less likely, given survival constraints and the rarity of monogenic vascular calcification syndromes. However, the absence of biochemical or histopathological data limits diagnostic certainty.

### MAC vs IAC: An Evolving Continuum

MAC and IAC have traditionally been considered distinct pathologies. However, emerging evidence supports a pathophysiological continuum shaped by systemic inflammation, phosphate dysregulation, vascular bed-specific factors, and mechanical stress (19, 20, 28, 58). Recent studies emphasize vascular smooth muscle cells (VSMCs) undergoing phenotypic switching toward an osteoblast-like state in response to pro-inflammatory and oxidative stressors, particularly under conditions of phosphate imbalance or oxidative stress, mediated primarily through NF-κB, RUNX2, BMP-2, and Wnt/β-catenin signaling pathways (15, 16, 18, 19, 59). MAC and IAC share these molecular pathways, suggesting common underlying mechanisms of vascular mineralization (20, 23, 58). This conceptual overlap is illustrated in Figure 4B.

In clinical populations (21) (21), this continuum is evident in overlapping calcification phenotypes among patients with chronic kidney disease (CKD) and diabetes mellitus, reflecting combined influences from vascular bed-specific mechanical factors, inflammatory stress, and phosphate metabolism dysregulation (3, 60–64). Epidemiological data emphasize that LDL cholesterol remains the predominant driver of IAC (65, 66), whereas MAC frequently arises in contexts involving phosphate imbalance and associated metabolic disturbances (8, 67). The calcifications observed in our 4400-year-old forearm specimen closely resemble those described in contemporary patients with chronic kidney disease, highlighting an evolutionarily conserved response to systemic metabolic or inflammatory stressors. This finding challenges a strict binary conceptualization of vascular calcification (IAC vs. MAC), reinforcing the need to acknowledge intermediate or transitional calcification forms and underscoring that arterial mineralization is not a purely modern phenomenon driven exclusively by lifestyle factors (23–26). Instead, vascular calcification appears to be a fundamental biological process conserved across historical contexts, responsive to metabolic and environmental stressors over millennia.

**Figure 4.**
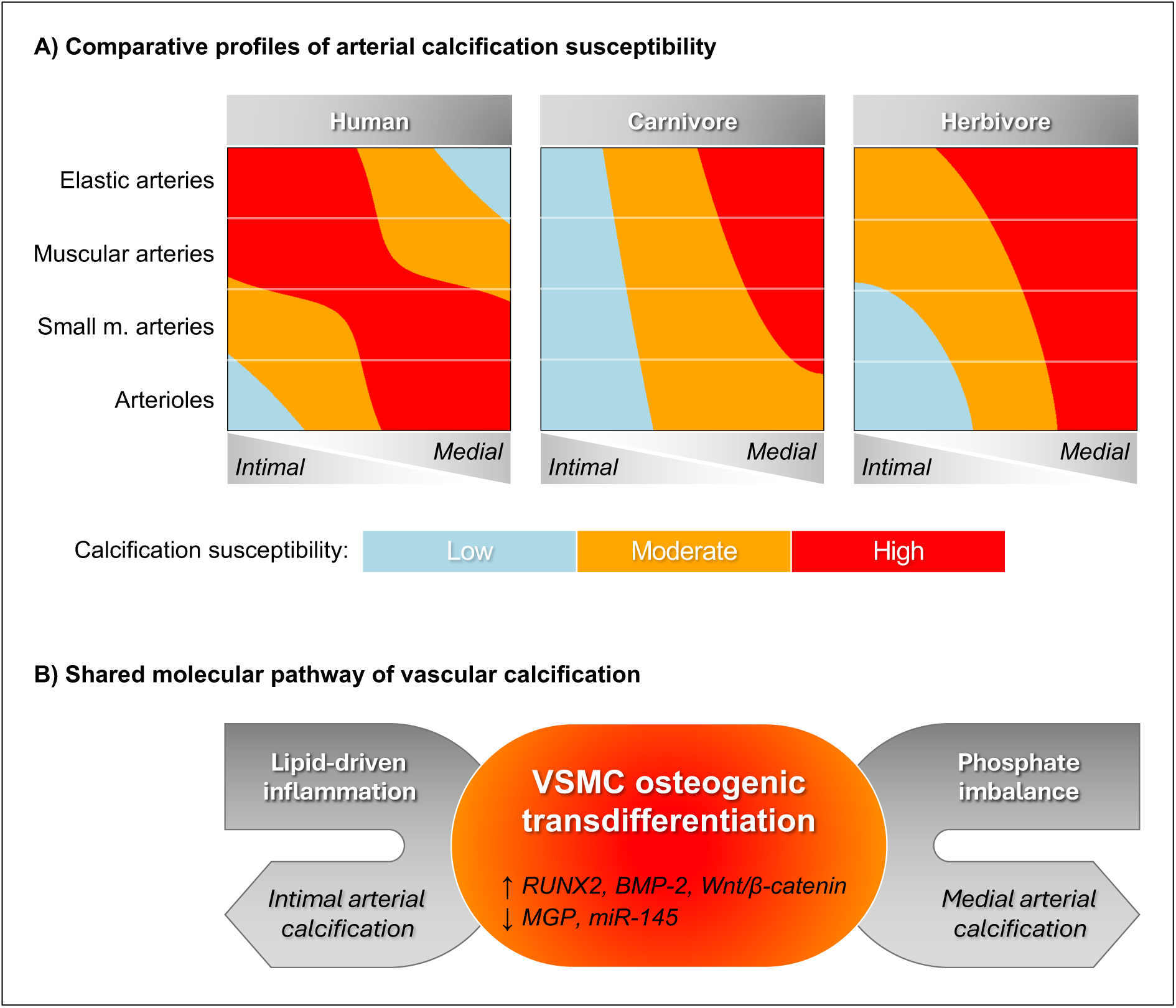
Conceptual landscape of arterial calcification susceptibility across representative vertebrate profiles. Panel A illustrates discrete viewports within a continuous, two-dimensional gradient space modeling arterial calcification susceptibility. Spatial axes represent artery type and size (vertical: distal to proximal; arterioles to elastic arteries) and arterial wall localization (horizontal: intimal to medial). The underlying gradient is color-coded into quantile-based susceptibility zones: low (blue), moderate (orange), and high (red). Each vertebrate profile occupies a distinct viewport, reflecting anatomical, physiological, and metabolic differences. Panel B summarizes shared osteogenic pathways implicated in intimal and medial calcification, including phosphate imbalance, vascular smooth muscle cell (VSMC) transdifferentiation, and inflammatory signaling.

### Evolutionary Lens and Cross-Species Observations

Paleopathological and veterinary studies indicate that vascular calcifications are neither uniquely modern nor confined to humans. Many nonhuman vertebrates exhibit arterial calcifications with highly variable, species-specific distributions (68–72), but animals–just like humans–can form intimal and medial calcifications, supporting the idea that vascular mineralization is an intrinsic but highly variable aspect of aging and physiological stress across taxa. Figure 4A illustrates how different vertebrate groups occupy distinct regions within a shared susceptibility landscape for vascular calcification.

Animal models, especially birds, show a more diffuse distribution of calcifications than humans (70). In domestic animals, primarily herbivores, the central arterial system is typically more affected by MAC than the smaller peripheral arteries. At the same time, IAC has been shown to affect herbivores only when fed large quantities of cholesterol or saturated fat (73). In contrast, in carnivores, experimental induction of atherosclerotic plaques has not been documented (72), while medial arterial calcifications are histologically similar to those in humans, albeit with considerable variability in their distribution patterns (71).

Despite extensive documentation of atherosclerotic lesions in mummies spanning diverse eras (e.g., the Horus Study) (23–26), MAC has rarely been explicitly categorized in ancient human remains. The identification of continuous rail-track calcifications in the distal upper limb arteries of a 4400-year-old specimen suggests that MAC may have historically been underrecognized, potentially overshadowed by focal intimal lesions at arterial branch points. Crucially, this also opens the possibility that distinctions between IAC and MAC may have been less pronounced in ancient populations than in contemporary clinical settings and did not necessarily hinge on established contemporary risk factors. Collectively, these insights underscore MAC as a longstanding biological response to chronic metabolic disturbances across diverse taxa, influenced by genetic, environmental, and physiological stressors, with phenotypic variability contingent upon species, population-specific conditions, and historical timeframes.

### Clinical and Translational Relevance

Identifying MAC in a 4400-year-old Old Kingdom Egyptian elite individual emphasizes its occurrence across vastly different environmental and dietary contexts, reinforcing MAC as an evolutionarily conserved pathology. This finding broadens the clinical perspective, underscoring the necessity of integrating metabolic, inflammatory, and infectious risk factors into contemporary diagnostic and therapeutic approaches to vascular calcification. Direct clinical implications arise from recognizing that MAC predates modern dietary and lifestyle factors, suggesting clinicians and researchers should consider broader, evolutionary consistent etiologies, including chronic infections, inflammation, and mineral dysregulation, particularly in atypical cases or resource-limited settings today.

#### Infection-Driven CKD as an Underexplored Contributor

The potential role of infection-driven CKD is particularly noteworthy, exemplified by ancient Egyptian populations with endemic schistosomiasis. Parasite-induced chronic renal impairment likely contributed significantly to phosphate dysregulation, a central driver of MAC today (8, 53, 55). Acknowledging infectious CKD as a historical contributor underscores the clinical importance of addressing infectious etiologies in contemporary healthcare, particularly in regions with ongoing high burdens of chronic infection and limited diagnostic capabilities.

#### Clinical Implications for Diagnostic Approaches

The conventional binary classification of vascular calcification as intimal (IAC) or medial (MAC) may oversimplify a complex underlying pathophysiological continuum. Recognizing transitional or mixed calcification forms, especially in contexts like CKD or diabetes, where patterns frequently overlap due to combined effects of vascular bed-specific mechanical stressors and systemic inflammatory conditions, could enhance diagnostic accuracy and risk stratification. Misclassification between MAC and IAC might represent a relevant clinical management pitfall, underscoring the importance of developing refined imaging protocols and novel biomarkers for improved differentiation (18, 19, 58).

#### Integration of Evolutionary Insights into Vascular Disease Research

This study illustrates the evolutionary persistence of MAC, underlining the importance of an evolutionary perspective in understanding vascular disease. Incorporating insights from paleopathology into contemporary vascular biology enhances our understanding of MAC pathogenesis as a fundamental adaptive response to chronic metabolic, inflammatory, and infectious stressors. This perspective supports the continuum hypothesis of arterial calcification. It may inform the identification of novel therapeutic targets, innovative preventive strategies, and enhanced approaches to clinical management, bridging historical insights with modern clinical practice.

### Limitations and Future Directions

This study is limited by its reliance on a single mummified forearm, restricting broader inferences about the prevalence and etiologies of MAC in Old Kingdom Egypt. While the findings provide compelling evidence for MAC in antiquity, a larger dataset is necessary to assess its epidemiological significance within ancient populations.

A further limitation is the lack of histological confirmation. The classification of calcifications as medial rather than intimal was based solely on planar radiography, which, while valuable for morphological assessment, does not provide direct evidence of tissue composition. If ethically and logistically feasible, clinical computed tomography, micro-CT, or limited histological sampling could confirm the precise layer of vascular mineralization and refine our understanding of vascular calcification patterns in ancient remains (74, 75).

Chronological precision is another challenge, as radiocarbon dating of Egyptian mummies is often subject to slight offsets due to embalming substances or dietary reservoir effects. While the dating results place the specimen within the expected Old Kingdom timeframe, additional isotopic analyses could further refine its historical context.

The etiology of MAC in this case remains somewhat speculative, as no biochemical analyses, parasite DNA studies, or molecular genetic testing were conducted. Although infection-driven CKD, metabolic dysregulation, and genetic predisposition are plausible contributors, confirming these hypotheses requires targeted biomolecular investigations. Future research should incorporate advanced imaging, stable isotope profiling of diet, and comparative analyses of additional mummies or ancient remains to expand our understanding of vascular calcification in antiquity.

Broadening the dataset to include multiple individuals from diverse periods and geographic regions would help clarify how environmental factors, infectious disease burden, and metabolic adaptations influenced arterial calcification across human history. Such investigations could bridge paleopathology with contemporary vascular biology, shedding light on the evolutionary drivers of vascular disease and refining modern perspectives on its pathogenesis.

## Conclusion

Our identification of probable MAC in an over 4,400-year-old mummified ancient Egyptian forearm underscores the longstanding and widespread nature of vascular mineralization processes. Although the absence of histological analysis precludes absolute confirmation, the continuous, tubular morphology of the calcifications strongly suggests MAC, rather than the patchy deposits characteristic of IAC, in a context long predating modern risk factors, such as high dietary lipids, widespread hypertension, and prolonged survival facilitated by insulin therapy.

Potential etiologic contributors, including chronic kidney disease secondary to endemic schistosomiasis, documented in Old Kingdom Egypt, and age-related degenerative or genetic factors, appear capable of producing morphologic outcomes similar to those seen in present-day CKD-related MAC. These findings challenge the notion that arterial calcification, regardless of its specific location in the vessel wall, is primarily a modern phenomenon, instead indicating a fundamental biological response that has endured for millennia.

Viewing MAC and IAC as a continuum of vascular pathology across time is not just of historical interest but carries modern implications. It suggests that clinicians should recognize overlaps between ‘atherosclerotic’ and ‘medial’ disease processes, especially in patients with atypical risk profiles (e.g., CKD or chronic inflammation). Our deep-time findings encourage an integrated view of vascular calcification, one that acknowledges how ancient trade-offs and exposures still shape disease expression today. In light of Thomas et al.’s commentary, which urges us to understand our evolutionary heritage (21), our study provides tangible evidence of that heritage – demonstrating that the seeds of cardiovascular disease were sown long ago, and continue to inform present-day pathology.

Detecting MAC in a single Old Kingdom individual may be a starting point for expanded paleopathological research and comparative studies. A better understanding of shared osteogenic pathways and longstanding environmental influences on vascular disease may inform novel risk stratification tools, personalized preventive strategies, and therapeutic interventions to mitigate IAC and MAC. Consequently, our paleopathological data not only sheds light on the health conditions of ancient Egyptians but also provides tangible insights that could influence how clinicians conceptualize and manage arterial calcification in contemporary populations.

## Data Availability

All data generated or analyzed during this study are included in the manuscript and its supplementary materials. Radiographic images are available upon reasonable request from the corresponding author, subject to institutional and curatorial permissions.

## Acknowledgments

We thank the Egyptian Museum in Cairo and its staff, particularly the Restoration Laboratory, for their support of this study, and Dr. Michael Habicht for his insightful recommendations on relevant literature.

## Author contributions

PE, FR, and MMO conceptualized the study. MMO, PE, and FR, supervised the research. PE, FR, and MMO administered the project. FR acquired funding for the research. PE, IH, and KDV developed the methodology. PE, IH, and MI carried out the investigations. PE, KDV, IH, and AEW, conducted formal analysis of the data. PE, IH, and AEW completed visualization. PE and KDV wrote the original draft of the manuscript. PE, KDV, IH, MI, and AEW were involved in reviewing and editing the manuscript.

